# Spatial inequality hides the burden of dog bites and the risk of dog-mediated human rabies

**DOI:** 10.1101/2020.02.07.20020727

**Authors:** Micaela De la Puente-León, Michael Z. Levy, Amparo M. Toledo, Sergio Recuenco, Julianna Shinnick, Ricardo Castillo-Neyra

## Abstract

Currently, there is an active rabies outbreak in the dog population of Arequipa, Peru. Inhabitants of the city are bitten by both pets and free-roaming dogs; therefore, the risk of human rabies transmission is concerning. Our objective was to estimate the rate of dog bites in the city and to identify factors associated with going or not going to a medical facility for rabies follow-up. To this end, we conducted a door-to-door survey of 4,000 houses in 21 urban and 21 peri-urban communities. We then analyzed associations between attaining follow-up rabies care and various socioeconomic factors, stratifying by urban and peri-urban localities. We found that the rate of dog bites in peri-urban communities (12.4%) was approximately three times higher than urban areas (4.0%). Among the people who were bitten, a significantly greater number of people in urban areas got follow-up rabies care than those in peri-urban areas.

## INTRODUCTION

Arequipa, an Andean city of approximately one million people, has grown continuously over the past six decades ^1 2^. The population growth of the city has been sustained by continuous migration, mostly from the countryside ^3^. The expansion of the city has occurred outwardly from the city center to the peripheral areas, as is the case in many South American cities ^1 4 5^. Therefore, a centrifugal urbanization gradient that differentiates urban and peri-urban areas in Arequipa can easily be observed in the urban landscape in Arequipa ^5^.

Rabies was reintroduced to the dog population of Arequipa in 2015 ^6,7^. Continuous transmission of the rabies virus, with one new case per week in 2019 indicates that the area is becoming endemic once again. The risk of acquiring rabies is influenced by exposure to a bite from a dog with rabies and access to treatment following exposure. 99% of human rabies deaths worldwide can be attributed to dog-mediated infections ^8^. Fortunately, human rabies has not been detected in Arequipa since 1990 ^9^. However, people still suffer dog bites in Arequipa and, therefore, there is potential for human rabies cases. Following the reintroduction of rabid dogs in 2015, the annual number of dog bites rose from 2,599 in 2014 to 6,621 in 2015. This increase is likely due to an increase in awareness and reporting following citywide efforts to disseminate information about rabies prevention. ^10 11^. Nonetheless, the number of bites not reported to the health system is unknown and is a threat to the rabies control program in Peru. Hundreds of rabid dogs have been detected since the beginning of the outbreak and virtually every district of the city has had at least one case ^12 13 14,15^.

Rabies is invariably a lethal disease, but it is also preventable. ^16 17^. The most effective strategy for human rabies prevention is to act on the animal reservoir, utilizing mass dog vaccinations to stop transmission ^18 19-21^. However, when people are bitten by dogs in areas with active transmission of the rabies virus, other preventative measures are necessary. In Arequipa and other dog-rabies endemic areas around the world, people must receive medical treatment when attacked by rabid reservoirs or potentially rabid animals.

Medical treatment for rabies includes a course of rabies vaccinations and wound management. Rabies post-exposure prophylaxis (PEP) are a series of vaccines provided when a person is attacked by rabid reservoirs or potentially rabid animals ^17,22^. PEP is highly effective at protecting those exposed to the rabies virus ^23^, but timely wound management is crucial ^8^. Wound management consists of cleaning the wound with soap and water, identifying the dog, and seeking attention at the nearest health facility; in some cases, anti-rabies serum might be required ^23,24^. Nearly 1 million people potentially exposed to rabies received PEP treatment annually just in the Americas ^25^. However, dog bite victims typically neglect treating their wounds appropriately, if at all, and many fail to seek medical treatment ^26 27 28 29^. In addition to the number of infected dogs, lack of access to appropriate medical treatment for rabies can also increase the risk of human rabies.

There are a greater number of free-roaming dogs in peri-urban than urban areas of Arequipa. Among these, there is a higher rate of rabid dogs detected in peri-urban areas that are more likely to bite and expose people to the virus. Given these two datapoints, we hypothesized that dog bites in peri-urban Arequipa may be underreported. Therefore, we conducted a door-to-door survey of 4,370 households in the Alto Selva Alegre (ASA) district of Arequipa, which spans both urban and peri-urban areas of the city. Our objectives were to estimate the dog bite rates in peri-urban and urban Arequipa and identify spatial and social factors associated with seeking healthcare following a dog bite incident. The results of this study provide information about spatial inequality in human rabies risk in Arequipa and insight into implementation of community strategies to improve both dog rabies surveillance and rabies healthcare access.

## 2. METHODOLOGY

### 2.1 Ethics statement

Ethical approval was obtained from Universidad Peruana Cayetano Heredia (approval number: 65369), Tulane University (approval number: 14–606720), and University of Pennsylvania (approval number: 823736).

### 2.2. Study setting

This study was conducted in the Alto Selva Alegre district (ASA) of Arequipa (human population in 2016: 83,310), one of the city’s 14 districts [GERSA AREQUIPA, 2018]. Arequipa, Peru’s second largest city, is home to 969,000 people. The city of Arequipa comprises communities spanning different stages of urbanization and different migration histories, from neighbourhoods established at the beginning of the 20th century, to recent developments ^5^. Within this gradient of development, most of the young neighbourhoods are located on the periphery of the city (peri-urban area) and the older localities are nearer to the center (urban area) ^5^. Compared to urban areas of the city, peri-urban areas generally have distinct geographic features, with rugged and uneven terrain (Figure 1). They are also marked by unique social characteristics, which include lower socioeconomic status, fewer community resources, and higher crime rates than urban areas. (Figure 1). As newer neighbourhoods mature into established neighbourhoods with wealthier residents, homes are improved with better quality construction material and permanent utility connections, and connectivity with the rest of the city increases with better sidewalks, roads, and transportation access. The ASA district transects the city, running from the center to the periphery and was therefore chosen as our study site because it encompasses both urban and peri-urban areas. Within the ASA district, the study sites include 21 urban neighborhoods founded multiple decades ago, and 21 peri-urban neighborhoods that originated around the year 2000 or later (Figure 1).

**Figure 1.**
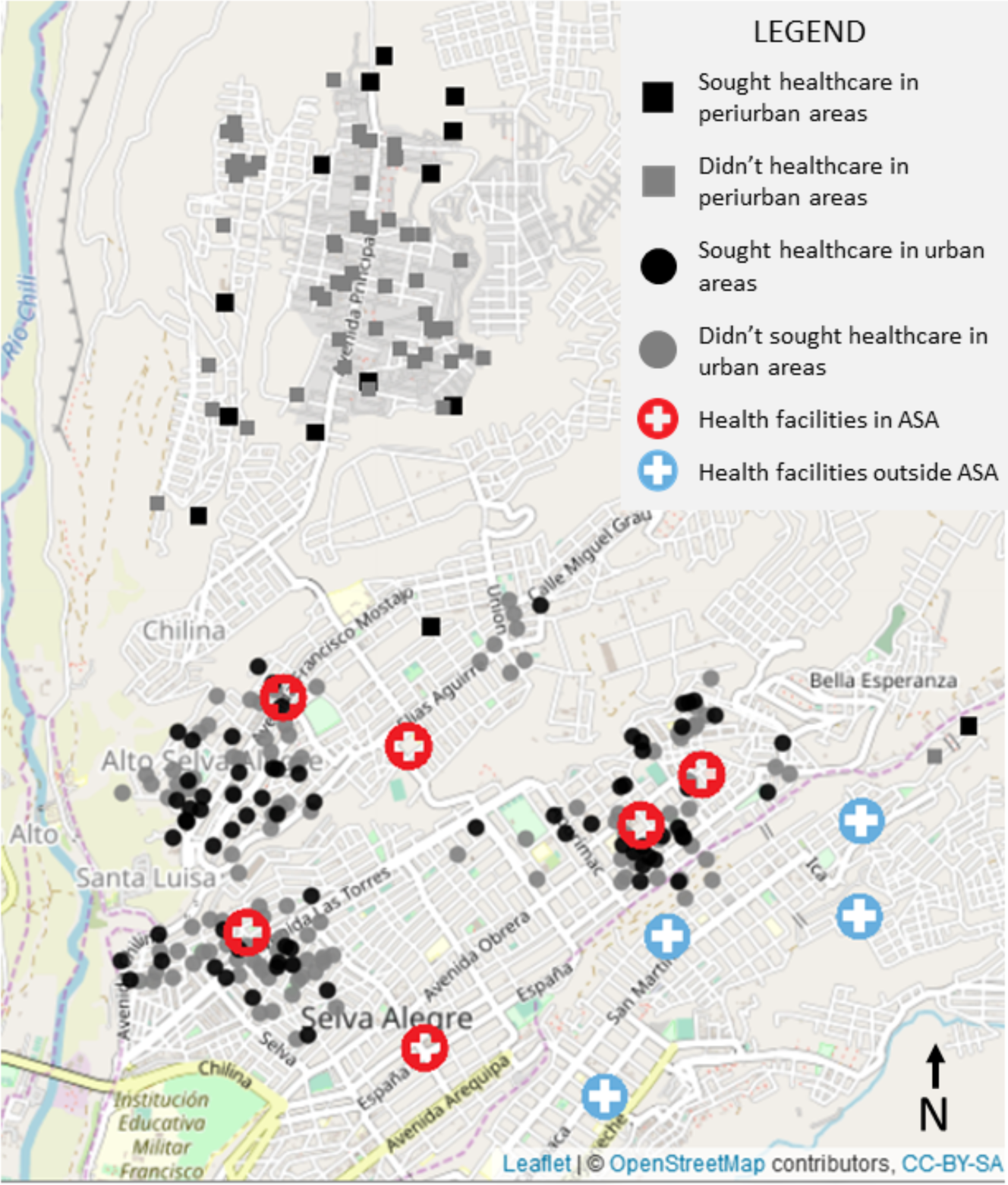
Map of bite cases and health facilities in urban versus peri-urban Arequipa. A higher proportion of adults from urban households sought healthcare following a dog bite (black). Real locations of the households have been modified using jitter procedure to protect confidentiality.

### 2.3 Data Collection

In September 2016, our team surveyed 4,370 households in the Alto Selva Alegre district in Arequipa. Surveys were performed door-to-door and lasted approximately 25 minutes. They contained questions regarding family composition of the household; dogs in the house; the interviewee’s socio-demographic information; dog bite incidents; and people’s actions following dog bite incidents. Each interview was of one adult, usually the person who answered the door. All houses were previously georeferenced by our team for other studies.

We also georeferenced all health posts and centers in Arequipa city. This includes 88 health facilities in total (30 health centers and 58 health posts), 6 of them are located in ASA (3 health centers and 3 health posts). No new health facilities have been built in ASA in the last decade ^30^ and there are no hospitals in the ASA district. We then calculated the shortest route from the households to the nearest health facility by foot. This approach assumed that community members would choose to go to the nearest public health facility and was not necessarily the actual route followed by those who sought care or the routes that those who did not seek care would have taken. Participants were asked if they sought care but not about the location of the health facility they visited; if it was public or private; or the level of the health facility (post, health center, clinic, hospital, etc).

The socio-demographic information we obtained from the dog bite survey included gender, age, and educational attainment. We also gathered household data including total number of people in the household, number of people under 18 years old (the age of majority in Peru), number of children under 5 years old, the interviewee’s length of residence in the house, and number of dogs living in the house. We also registered the ownership status (own or unknown dog) and vaccination status of the dog that bit the participant. If the person did not know the vaccination status of the dog that bit them, we considered the dog to not be vaccinated, as is standard in local and international procedures to apply PEP in those cases 24,31.

### 2.4. Statistical Analysis

We collected data from bitten adults only, as they are capable of making a decision about whether or not to seek medical care. We also calculated the shortest distance from the house to the nearest health facility using Leaflet, an open-source JavaScript library for interactive maps that can calculate routes by car, bike, public transportation and walking ^32^. We set each bitten person’s house as a starting point and each health facility in Arequipa as an endpoint, via Leaflet Routing Machine services and Mapbox Directions API ^33,34^, and then we calculated the shortest walking distance.

We explored the spatial distribution of the bite cases and health facilities in urban and peri-urban areas. To evaluate the characteristics of interviewees, we first compared those who were bitten with those who were not (Table 1). Secondly, among those who were bitten, we compared the baseline characteristics of those who sought healthcare with those who did not, stratifying by type of locality (urban and peri-urban) (Table 2). For both analyses, we used a chi-square test to compare categorical variables with 10 or more observations per group, Fisher’s exact test for categorical variables with fewer than 10 observations in any subgroup, and Mann–Whitney U test for age of the dog owner or interviewee, which did not follow a normal distribution and was truncated at 18 years. We also evaluated correlations between our continuous variables by type of locality. For distance to a healthcare facility, we used the shortest walking distance from the house to the closest healthcare facility, either health center or health post.

**Table 1.** Descriptive statistics of associated variables with being bitten or not being bitten by a dog in Alto Selva Alegre district in Arequipa, 2016. All confidence intervals were calculated at 95% * Mann-Whitney U-test & T-test £ Chi-square test $ Fisher test

|  | Bitten by a dog<br>(n=254) | Not bitten by a<br>dog (n=4016) | p-value |  |
| --- | --- | --- | --- | --- |
| Age (IQR) | 38 (26 - 53) | 40 (28 - 53) | 0.242 | * |
| Mean age (SD) | 40.49 (16.2) | 41.61 (15.9) | 0.282 | & |
| Females % | 61.8 | 66.8 | 0.026 | £ |
| Instruction level | | | 0.725 | \$ |
| Primary school (%) | 13.4 | 12.3 |  |  |
| Secondary school (%) | 40.6 | 42.4 |  |  |
| Technical school (%) | 20.1 | 21.3 |  |  |
| University (%) | 25.2 | 23.5 |  |  |
| Illiterate (%) | 0.8 | 0.5 |  |  |
| Median number of people in the house (IQR) | 4 (3 - 6) | 4 (3 - 6) | 0.407 | * |
| Mean number of people in the house (CI) | 4.49 (4.23-4.75) | 4.63 (4.56-4.71) | 0.293 | & |
| Median number of people < 18yo (IQR) | 0 (0 - 1) | 0 (0 - 1) | 0.712 | * |
| Mean number of people < 18yo (CI) | 0.78 (0.71-0.85) | 0.78 (0.75-0.81) | 0.962 | & |
| Median number of children < 5yo (IQR) | 0 (0 - 1) | 0 (0 - 1) | 0.947 | * |
| Mean number of children < 5yo (CI) | 0.48 (0.38-0.57) | 0.47 (0.45-0.5) | 0.965 | & |
| Median length of residence (year) (IQR) | 15 (5 - 25) | 16 (6 - 26) | 0.356 | * |
| Has dogs (%) | 66.1 | 70.9 | 0.03 | £ |
| Median number of dogs (IQR) | 1 (0 - 2) | 1 (0 - 2) | 0.213 | * |
| Mean number of dogs (CI) | 1.35 (1.18-1.52) | 1.24 (1.2-1.27) | 0.195 | & |

**Table 2.** Descriptive statistics of associated variables to type locality in Alto Selva Alegre district in Arequipa, 2016.

|  | Peri-urban (n=70) | Urban (n=184) | p-value |  |
| --- | --- | --- | --- | --- |
| Median shortest distance (IQR) | 2439.6 (2224.7 - 2771.9) | 398.4 (252.8 - 618.4) | < 0.001 | * |
| Mean shortest distance (sd) | 2427.2 (436.3) | 448.5 (233) | < 0.001 | & |
| Age (IQR) | 34 (26 - 43) | 40 (27 - 56) | 0.02 | * |
| Mean age (sd) | 36.4 (14.1) | 42.0 (16.7) | 0.008 | & |
| Females % | 64.3 | 62 | 0.843 | £ |
| Educational attainment |  |  |  |  |
| *Primary school (%)* | 24.3 | 9.2 | 0.001 | \$ |
| *Secondary school (%)* | 47.1 | 38 |  |  |
| *Technical school (%)* | 15.7 | 21.7 |  |  |
| *University (%)* | 11.4 | 30.4 |  |  |
| *Illiterate (%)* | 1.4 | 0.5 |  |  |
| Median number of people in the house (IQR) | 4 (3 - 5) | 4 (3 - 6) | 0.007 | * |
| Mean number of people in the house (CI) | 3.83 (3.5-4.16) | 4.74 (4.41-5.08) | < 0.001 | & |
| Median number of people < 18yo | 1 (0 - 1.75) | 0 (0 - 1) | 0.175 | * |
| (IQR) |  |  |  |  |
| Mean number of people < 18yo (CI) | 0.91 (0.68-1.15) | 0.73 (0.6-0.86) | 0.191 | & |
| Median number of children < 5yo (IQR) | 0 (0 - 1) | 0 (0 - 1) | 0.609 | * |
| Mean number of people < 5yo (CI) | 0.46 (0.32-0.6) | 0.48 (0.37-0.6) | 0.783 | & |
| Median length of residence (year) (IQR) | 7 (3 - 10) | 20 (9.5 - 30) | < 0.001 | * |
| Owns dogs (%) | 81.4 | 64.7 | 0.015 | £ |
| Median number of dogs (IQR) | 1.5 (1 - 2.75) | 1 (0 - 2) | 0.003 | * |
| Mean number of dogs (CI) | 1.74 (1.37-2.12) | 1.2 (1.02-1.38) | 0.013 | & |
| Bitten by own dog (%) | 15.7 | 10.9 | 0.401 | £ |
| Bitten by vaccinated dog (%) | 20 | 31.1 | 0.108 | £ |
|  | Peri-urban (n=70) | Urban (n=184) | p-value |  |
| Median shortest distance (IQR) | 2439.6 (2224.7 - 2771.9) | 398.4 (252.8 - 618.4) | < 0.001 | * |
| Mean shortest distance (sd) | 2427.2 (436.3) | 448.5 (233) | < 0.001 | & |
| Age (IQR) | 34 (26 - 43) | 40 (27 - 56) | 0.02 | * |
| Mean age (sd) | 36.4 (14.1) | 42.0 (16.7) | 0.008 | & |
| Females % | 64.3 | 62 | 0.843 | £ |
| Educational attainment |  |  |  |  |
| *Primary school (%)* | 24.3 | 9.2 | 0.001 | \$ |
| *Secondary school (%)* | 47.1 | 38 |  |  |
| *Technical school (%)* | 15.7 | 21.7 |  |  |
| *University (%)* | 11.4 | 30.4 |  |  |
| *Illiterate (%)* | 1.4 | 0.5 |  |  |
| Median number of people in the house (IQR) | 4 (3 - 5) | 4 (3 - 6) | 0.007 | * |
| Mean number of people in the house (CI) | 3.83 (3.5-4.16) | 4.74 (4.41-5.08) | < 0.001 | & |
| Median number of people < 18yo (IQR) | 1 (0 - 1.75) | 0 (0 - 1) | 0.175 | * |
| Mean number of people < 18yo (CI) | 0.91 (0.68-1.15) | 0.73 (0.6-0.86) | 0.191 | & |
| Median number of children < 5yo (IQR) | 0 (0 - 1) | 0 (0 - 1) | 0.609 | * |
| Mean number of people < 5yo (CI) | 0.46 (0.32-0.6) | 0.48 (0.37-0.6) | 0.783 | & |
| Median length of residence (year) (IQR) | 7 (3 - 10) | 20 (9.5 - 30) | < 0.001 | * |
| Owns dogs (%) | 81.4 | 64.7 | 0.015 | £ |
| Median number of dogs (IQR) | 1.5 (1 - 2.75) | 1 (0 - 2) | 0.003 | * |
| Mean number of dogs (CI) | 1.74 (1.37-2.12) | 1.2 (1.02-1.38) | 0.013 | & |
| Bitten by own dog (%) | 15.7 | 10.9 | 0.401 | £ |
| Bitten by vaccinated dog (%) | 20 | 31.1 | 0.108 | £ |

Our main objective was to identify spatial and social factors associated to seeking healthcare following a dog bite incident. To describe responses to dog bite incidents, we used categorical values: sought healthcare and did not seek healthcare. We ran univariable and multivariable logistic regression to test variables previously found to be associated with seeking healthcare ^27,28,35-37 38-42^. Based on our univariable analysis, the following covariates were used to build the model: amount of time living in the house, whether the bite was from the participant’s own dog, distance to closest healthcare facility, gender of the participant, and educational attainment. However, we also explored models via stepwise forward and back selection including all the variables evaluated. We also applied principal component analysis to several variables including distance to the closest healthcare facility, length of residence in the house, and age of the interviewee to reduce the dimensionality of the data while accounting for the collinearity. All data management and analysis were performed in R version 3.3.1 -© 2016, Inc.

## RESULTS

We visited 6,420 houses during the survey period. 4,370 community members (each for a unique household) consented to participate in the study and were surveyed. 254 interviewees (5.8%, 95% CI: 5.2 to 6.5) reported suffering a dog bite in the last 12 months: 184 in urban areas and 70 in peri-urban areas (Figure 1). The annual dog bite rate was 4.96% in urban areas and 12.43% in peri-urban areas.

The probability of suffering a dog bite was higher in women and people with at least one dog at home. Neither age of the participants (all adults), educational attainment, nor number of dogs at home were associated with suffering a dog bite. Other evaluated variables can be found in Table 1.

Among those who suffered dog bites in urban areas, 39.1% sought medical care at a health post, center or hospital after the bite incident, compared to 21.4% in peri-urban areas. (overall medical seeking behavior = 65.7%, 95% CI: 60.0 to 71.3). Variables associated with receiving medical attention after a dog bite were age of bitten adults, shortest-route distances to the closest health post or center, number of years living in the house, number of dogs in the house and ownership of the dog that bit them (Figure 2). The probability of receiving medical attention decreased with greater distances between the house and the nearest health facility. A lower rate of seeking medical care was also associated with being bitten by one’s own dogs rather than unknown dogs. A higher probability of seeking medical attention was associated with a longer length of residence in the house and older age. Contrary to our expectations, we did not find an association between the vaccination status of the biting dog and seeking medical care. Association with the number of dogs in the house is unclear. Further information can be found in Table 2.

**Supplementary table 1.**
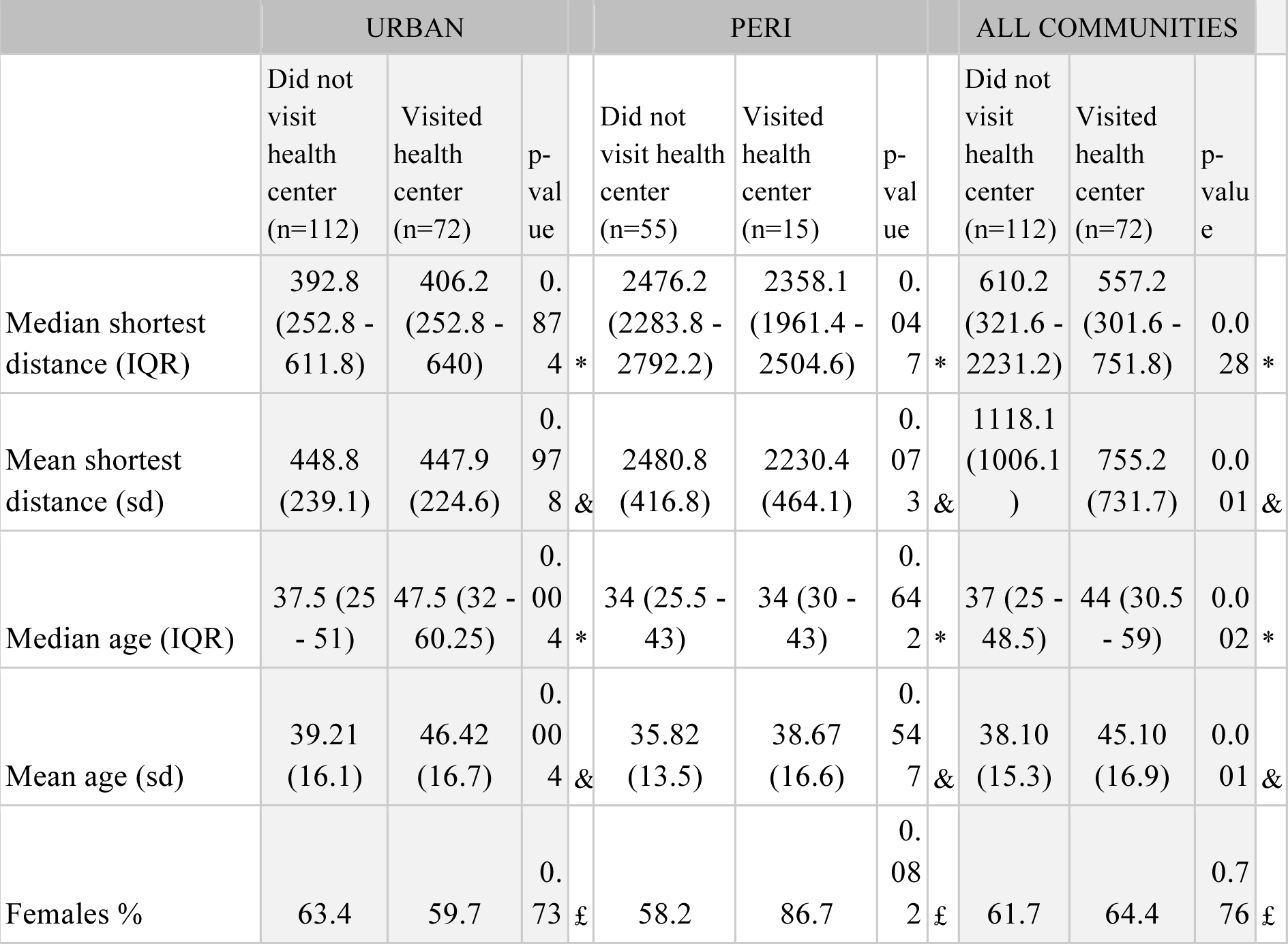
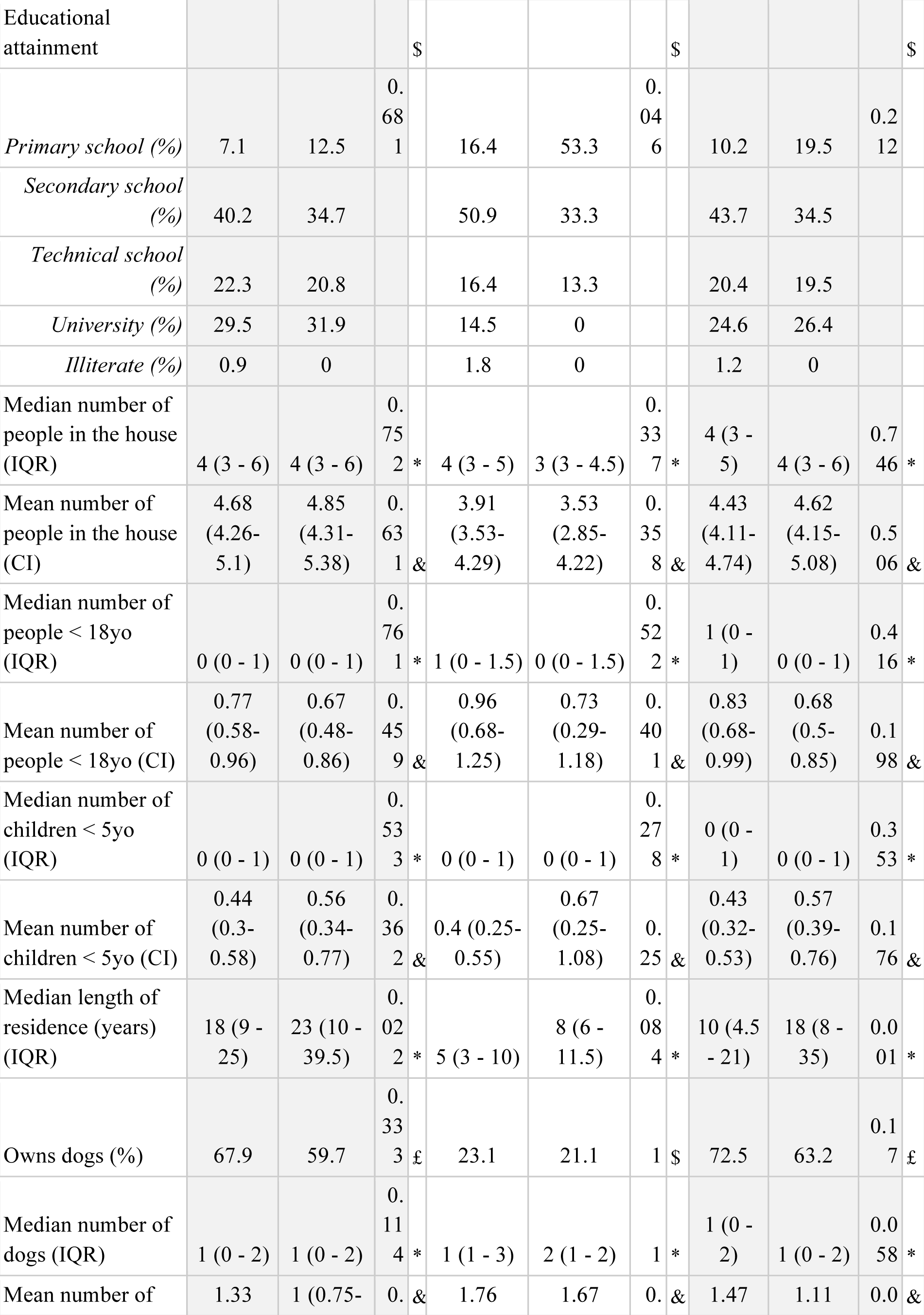
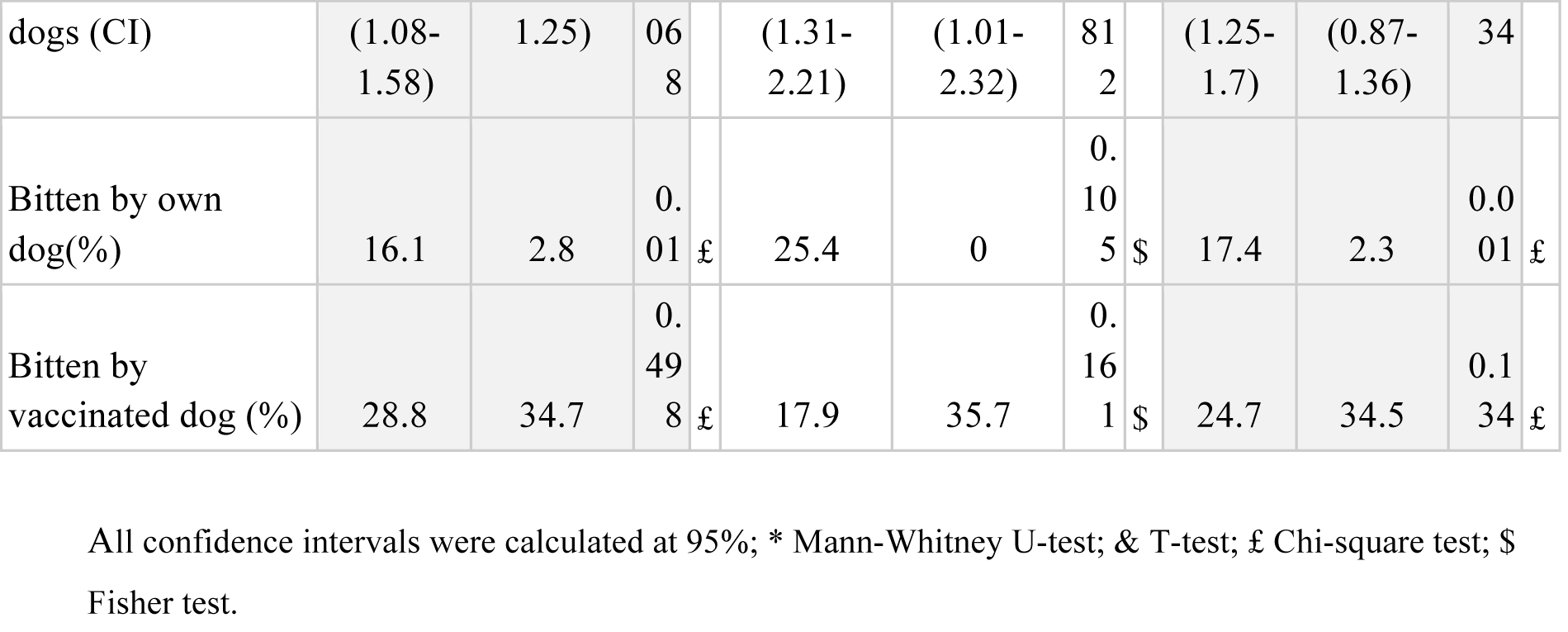
Descriptive statistics of associated variables with adults seeking healthcare or not after a dog bite incident in Alto Selva Alegre district in Arequipa, 2016.

**Figure 2.**
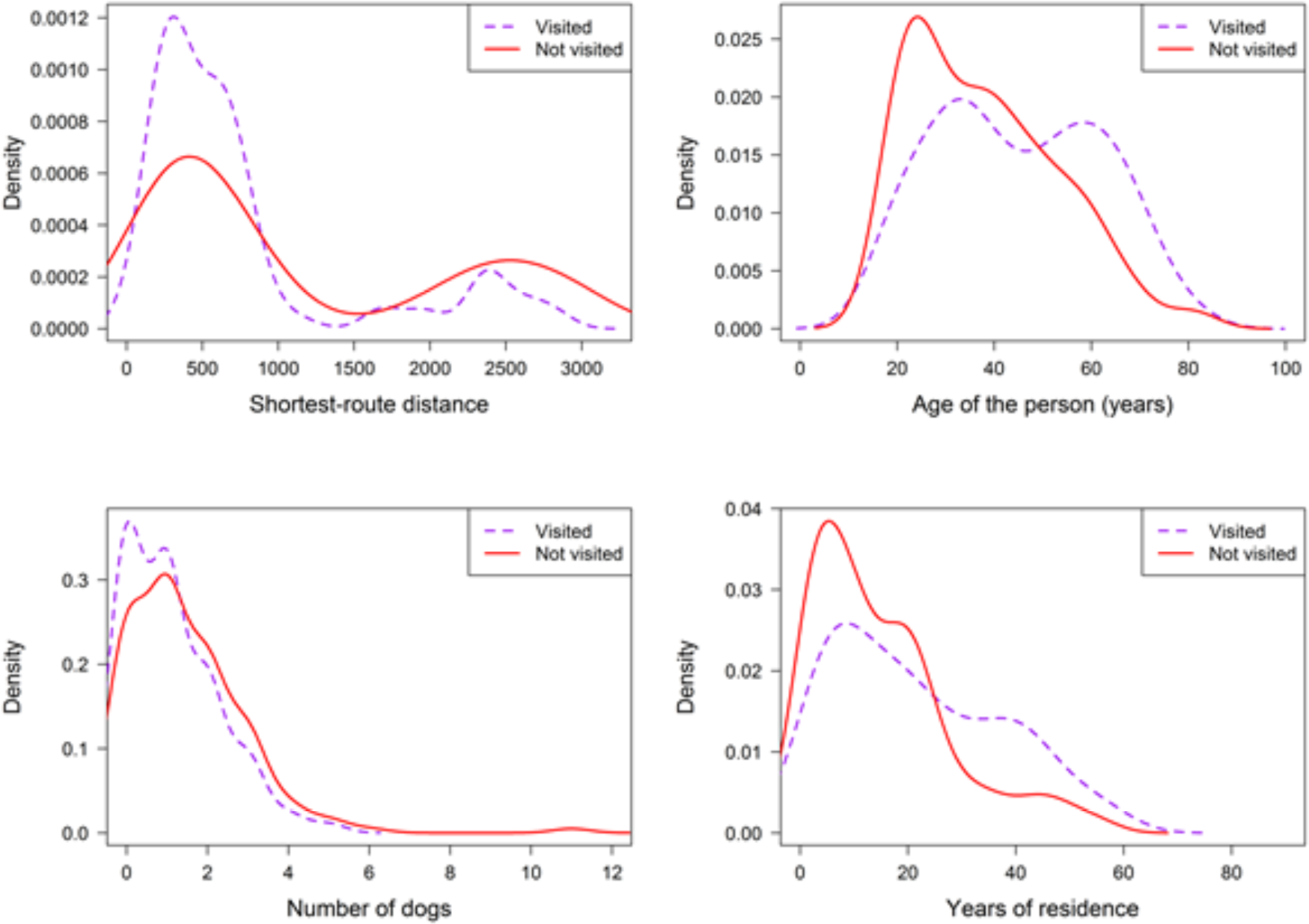
Density plot of people that visited or not visited a health facility following a dog bite incident, showing distribution of a) shortest-route distance in meters, b) age of the person in years, c) number of dogs in the house and d) length of residence (years) in the house.

All 6 health facilities in ASA were located in the urban communities of ASA; there were no health facilities in the peri-urban area (Figure 1). Walking distance to the nearest health center or health post ranged from a few meters to 3.2 km. A single health facility (C.S. Independencia) was the closest health facility for nearly half of the bitten adults (47.2%, 120/254). For some participants, the closest health facility was in a different district. We found several notable differences between the characteristics of the households in urban and peri-urban areas (Fig. 3). The distance from surveyed households to the nearest health post or center was significantly longer in peri-urban than urban areas. The number of people living in a household as well of length of residence was higher in urban areas. Residents in peri-urban areas were more likely to have a dog at home and owned a higher number of dogs on average than residents in urban areas. Other variables associated with peri-urban versus urban households in ASA are detailed in Table 3.

**Table 3.**
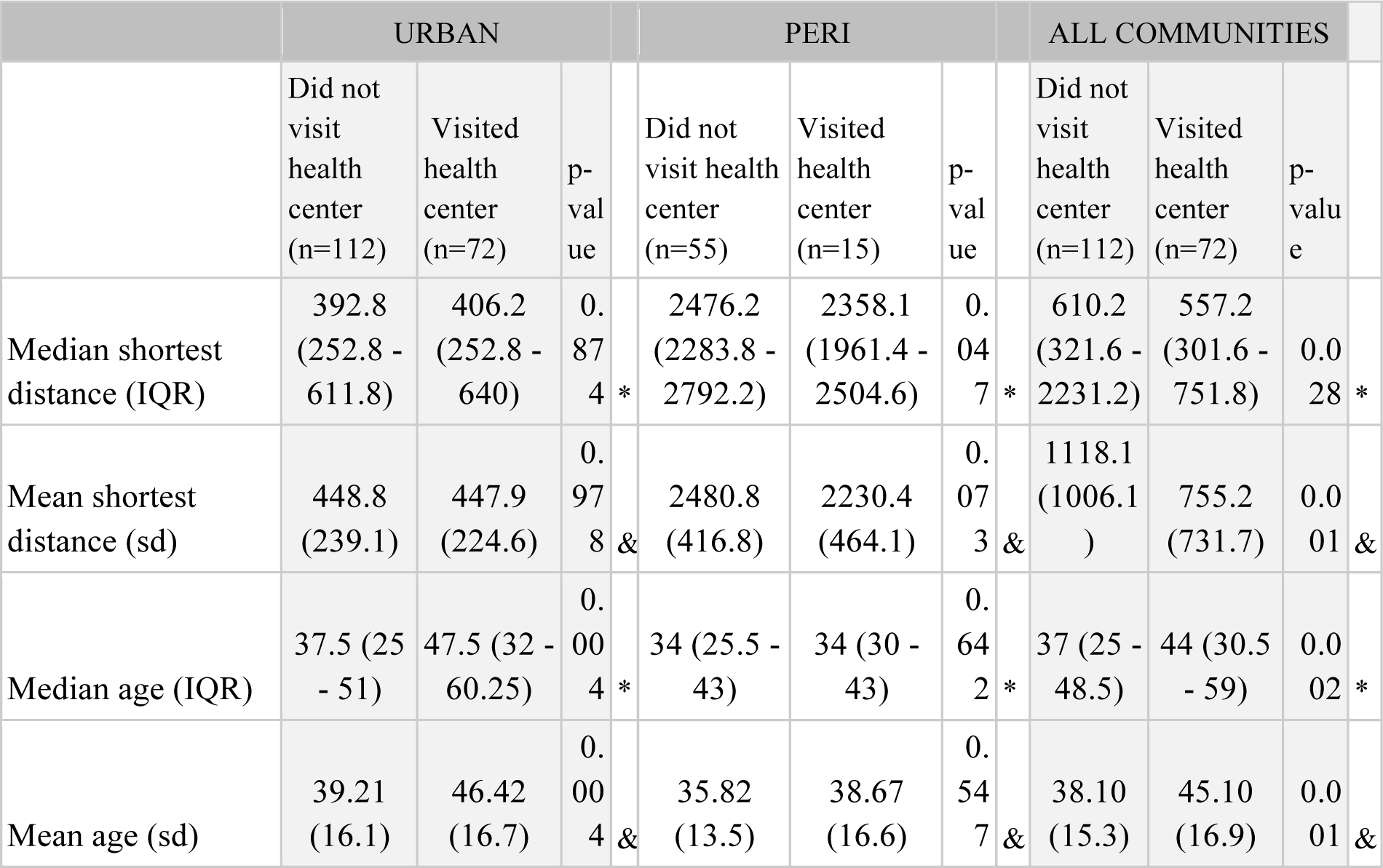
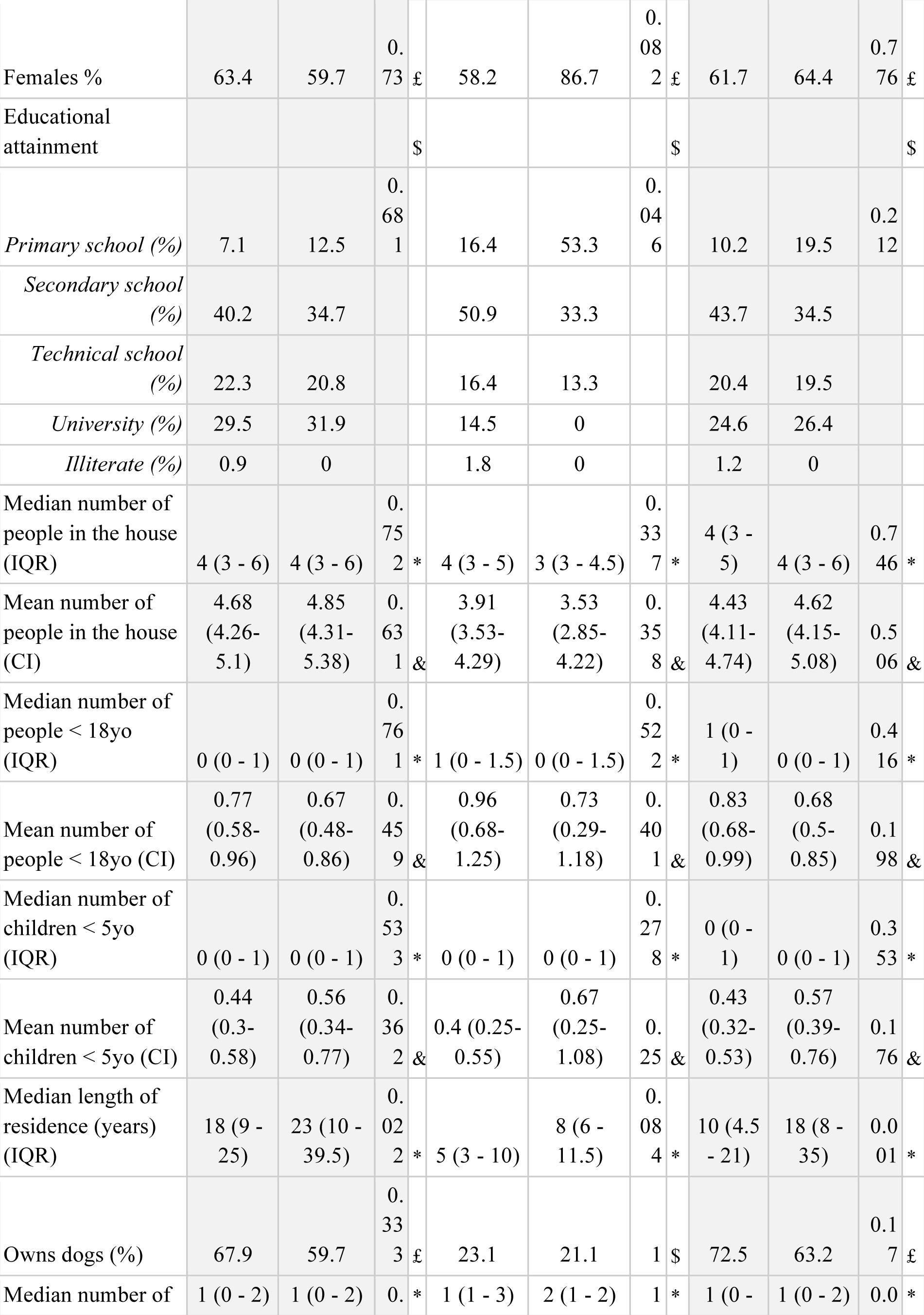
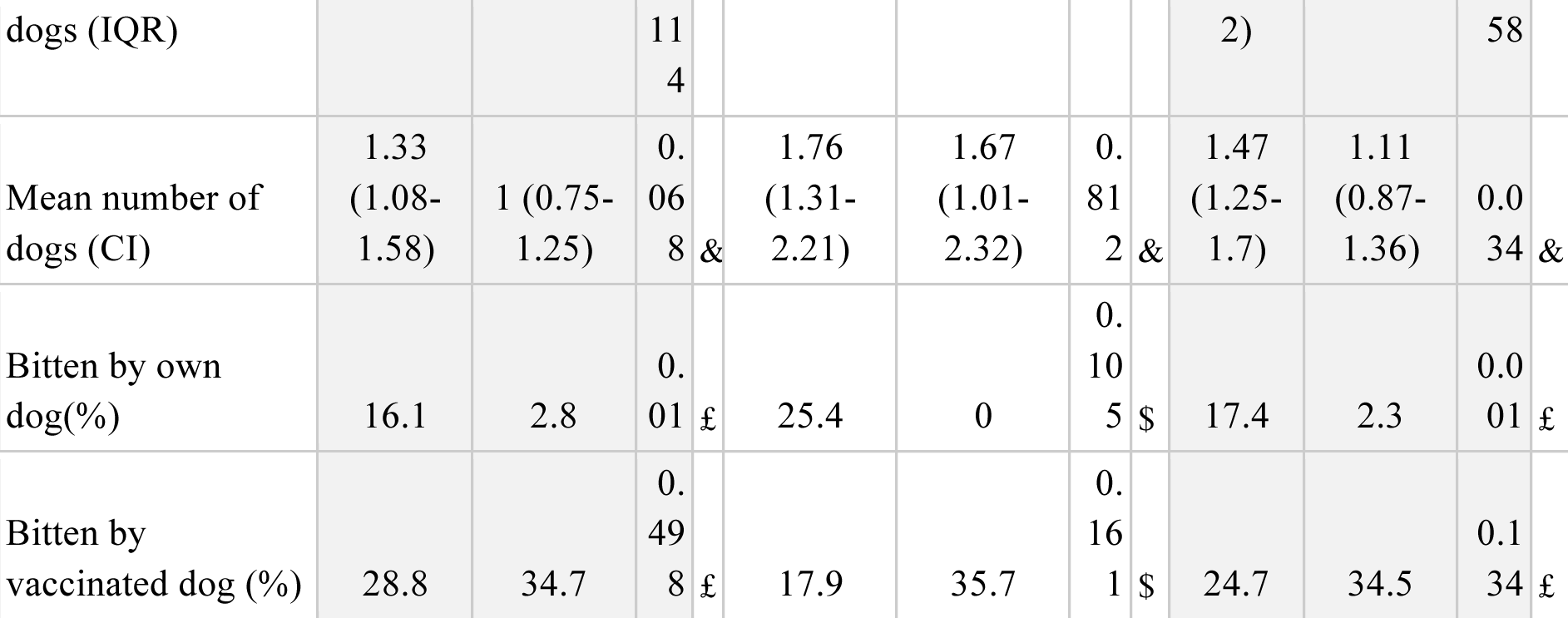
Best fitting model selected by stepwise procedure for adults seeking medical attention after a dog bite in all the communities, urban communities and peri-urban communities in Alto Selva Alegre district in Arequipa, 2016.

|  | URBAN |  |  | PERI |  |  | ALL COMMUNITIES |  |  |
| --- | --- | --- | --- | --- | --- | --- | --- | --- | --- |
|  | Did not visit health center (n=112) | Visited health center (n=72) | p-value | Did not visit health center (n=55) | Visited health center (n=15) | p-value | Did not visit health center (n=112) | Visited health center (n=72) | p-value |
| Median shortest distance (IQR) | 392.8<br>(252.8 - 611.8) | 406.2<br>(252.8 - 640) | 0.874 * | 2476.2<br>(2283.8 - 2792.2) | 2358.1<br>(1961.4 - 2504.6) | 0.047 * | 610.2<br>(321.6 - 2231.2) | 557.2<br>(301.6 - 751.8) | 0.028 * |
| Mean shortest distance (sd) | 448.8<br>(239.1) | 447.9<br>(224.6) | 0.978 & | 2480.8<br>(416.8) | 2230.4<br>(464.1) | 0.073 & | 1118.1<br>(1006.1 ) | 755.2<br>(731.7) | 0.001 & |
| Median age (IQR) | 37.5 (25 - 51) | 47.5 (32 - 60.25) | 0.004 * | 34 (25.5 - 43) | 34 (30 - 43) | 0.642 * | 37 (25 - 48.5) | 44 (30.5 - 59) | 0.002 * |
| Mean age (sd) | 39.21<br>(16.1) | 46.42<br>(16.7) | 0.004 & | 35.82<br>(13.5) | 38.67<br>(16.6) | 0.547 & | 38.10<br>(15.3) | 45.10<br>(16.9) | 0.001 & |

|  |  |  |  |  |  |  |  |  |  |  |  |  |
| --- | --- | --- | --- | --- | --- | --- | --- | --- | --- | --- | --- | --- |
| Females % | 63.4 | 59.7 | 0.73 | £ | 58.2 | 86.7 | 0.082 | £ | 61.7 | 64.4 | 0.776 | £ |
| Educational attainment | | | | \$ | | | | \$ | | | | \$ |
| *Primary school (%)* | 7.1 | 12.5 | 0.681 |  | 16.4 | 53.3 | 0.046 |  | 10.2 | 19.5 | 0.212 |  |
| *Secondary school (%)* | 40.2 | 34.7 |  |  | 50.9 | 33.3 |  |  | 43.7 | 34.5 |  |  |
| *Technical school (%)* | 22.3 | 20.8 |  |  | 16.4 | 13.3 |  |  | 20.4 | 19.5 |  |  |
| *University (%)* | 29.5 | 31.9 |  |  | 14.5 | 0 |  |  | 24.6 | 26.4 |  |  |
| *Illiterate (%)* | 0.9 | 0 |  |  | 1.8 | 0 |  |  | 1.2 | 0 |  |  |
| Median number of people in the house (IQR) | 4 (3 - 6) | 4 (3 - 6) | 0.752 | * | 4 (3 - 5) | 3 (3 - 4.5) | 0.337 | * | 4 (3 - 5) | 4 (3 - 6) | 0.746 | * |
| Mean number of people in the house (CI) | 4.68 (4.26-5.1) | 4.85 (4.31-5.38) | 0.631 | & | 3.91 (3.53-4.29) | 3.53 (2.85-4.22) | 0.358 | & | 4.43 (4.11-4.74) | 4.62 (4.15-5.08) | 0.506 | & |
| Median number of people < 18yo (IQR) | 0 (0 - 1) | 0 (0 - 1) | 0.761 | * | 1 (0 - 1.5) | 0 (0 - 1.5) | 0.522 | * | 1 (0 - 1) | 0 (0 - 1) | 0.416 | * |
| Mean number of people < 18yo (CI) | 0.77 (0.58-0.96) | 0.67 (0.48-0.86) | 0.459 | & | 0.96 (0.68-1.25) | 0.73 (0.29-1.18) | 0.401 | & | 0.83 (0.68-0.99) | 0.68 (0.5-0.85) | 0.198 | & |
| Median number of children < 5yo (IQR) | 0 (0 - 1) | 0 (0 - 1) | 0.533 | * | 0 (0 - 1) | 0 (0 - 1) | 0.278 | * | 0 (0 - 1) | 0 (0 - 1) | 0.353 | * |
| Mean number of children < 5yo (CI) | 0.44 (0.3-0.58) | 0.56 (0.34-0.77) | 0.362 | & | 0.4 (0.25-0.55) | 0.67 (0.25-1.08) | 0.25 | & | 0.43 (0.32-0.53) | 0.57 (0.39-0.76) | 0.176 | & |
| Median length of residence (years) (IQR) | 18 (9 - 25) | 23 (10 - 39.5) | 0.022 | * | 5 (3 - 10) | 8 (6 - 11.5) | 0.084 | * | 10 (4.5 - 21) | 18 (8 - 35) | 0.001 | * |
| Owns dogs (%) | 67.9 | 59.7 | 0.333 | £ | 23.1 | 21.1 | 1 | \$ | 72.5 | 63.2 | 0.17 | £ |
| Median number of | 1 (0 - 2) | 1 (0 - 2) | 0. | * | 1 (1 - 3) | 2 (1 - 2) | 1 | * | 1 (0 - | 1 (0 - 2) | 0.0 | * |

|  |  |  |  |  |  |  |  |  |  |  |
| --- | --- | --- | --- | --- | --- | --- | --- | --- | --- | --- |
| dogs (IQR) |  |  | 11<br>4 |  |  |  | 2) |  | 58 |  |
| Mean number of dogs (CI) | 1.33<br>(1.08-1.58) | 1 (0.75-1.25) | 0.06<br>8 & | 1.76<br>(1.31-2.21) | 1.67<br>(1.01-2.32) | 0.81<br>2 & | 1.47<br>(1.25-1.7) | 1.11<br>(0.87-1.36) | 0.0<br>34 & |  |
| Bitten by own dog(%) | 16.1 | 2.8 | 0.01<br>£ | 25.4 | 0 | 0.10<br>5 \$ | 17.4 | 2.3 | 0.0<br>01 £ | |
| Bitten by vaccinated dog (%) | 28.8 | 34.7 | 0.49<br>8 £ | 17.9 | 35.7 | 0.16<br>1 \$ | 24.7 | 34.5 | 0.1<br>34 £ | |

**Fig 3.**
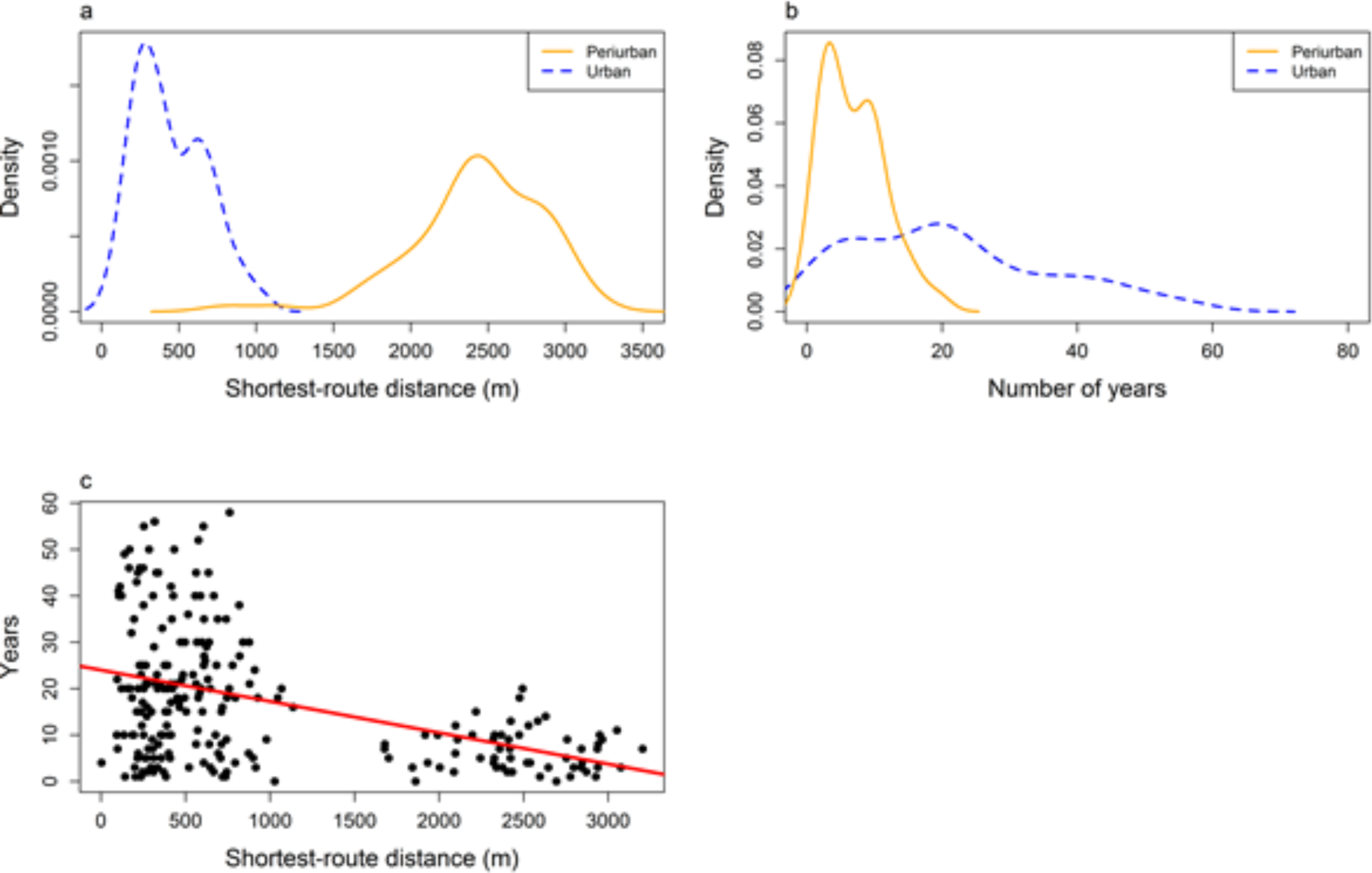
Correlation between the type of area with a) the shortest-route distance in meters from the house to the nearest health facility and b) length of residence.

Fig 3. Correlation between the type of area with a) the shortest-route distance in meters from the house to the nearest health facility and b) length of residence.

There was a significant positive correlation between distance from the house to the nearest health center and length of residence (p value < 0.005, cor. 0.44). As expected, there was a low to moderate positive correlation between the age of the surveyed person and length of residence (0.34) and moderate positive correlation between the total number of people and number of children under 18 years old living in house (0.41) and children under 5 years old living in the house (0.5). In addition, weak negative correlation was found between distance to the nearest health center and total number of people (0.21) and between number of children under 18 years old and children under 5 years old living in the house (0.21).

The best fitting model via forward and backward stepwise selection for all communities included the variables of interviewee’s age, length of residence and number of children under 5 years old in the house and ownership and vaccination status of the dog the bit (Table 4). However, the set of statistically significant variables was different when the model was stratified by urban and peri-urban areas. For urban communities, only length of residence in the house and ownership of the dog that bit remained significant. In contrast, in peri-urban communities, neither length of residence in the houses, distance to the closest healthcare facility, nor vaccination status of the dog that bit remained significant. Detailed results of the best fitting model are in Table 4.

## DISCUSSION

12.4% of people in peri-urban areas of ASA suffered a dog bite in the past year as compared with 4.0% of people living in urban areas of the district. This finding represents a clear spatial disparity in dog bite burden: there is an approximately three times greater risk of being bitten in a peri-urban area than an urban area of Arequipa. The higher rate of dog bites can be partially attributed to a greater density of street dogs in peri-urban areas than urban areas of the city ^40^. There are several geographic and social influences on street dog density in peri-urban areas. Geographically, there is more space for free-roaming dogs than in densely-populated urban areas. Torrenteras run through the peri-urban areas of Arequipa and have been previously described as “highways” for street dogs to traverse the city ^43^. Houses in peri-urban areas are also built with cheaper materials than urban dwellings. Characteristic stone or brick fences and corrugated metal roofs are not always effective at keeping pets inside or stray dogs out, especially if a female pet is in heat ^40,44^. Geographic characteristics of peri-urban Arequipa are partially responsible for a higher dog density in peri-urban areas.

There are also unique social influences on dog density, which stem from the poverty of peri-urban areas. The cost of neutering or spaying a dog is significant, particularly in peri-urban areas where residents have a lower average household income ^44^. When pet dogs have puppies, those puppies may be left in the streets if they are not sold or adopted. Residents of peri-urban areas may keep more dogs as protection due to higher rates of crime ^40^. Both geographic and social factors influence a higher density of street dogs, a higher rate of dog bites and, therefore, a spatial disparity in human rabies risk across the city.

The high rate of dog bites in peri-urban areas is especially concerning for rabies risk because dogs are less likely to be vaccinated against rabies in peri-urban than urban areas ^44,45^. Distance to vaccination points and rugged topography of peri-urban regions are barriers for community members to vaccinate their dogs ^44,45^. This finding suggests that a “one health” approach to rabies prevention in peri-urban Arequipa, which addresses rabies prevention in pets, wild animals and humans may lead to more effective interventions to decrease spatial inequality. The large population of unvaccinated dogs on the streets and a high rate of dog bites influences greater rabies risk in peri-urban areas.

Once bitten, fewer people in peri-urban areas than urban areas of Arequipa receive medical treatment, continuing a trend of spatial inequality in human rabies risk. Follow-up treatment for dog bites is essential in decreasing human rabies risk because the healthcare professionals can clean the wound, decreasing viral load, and can provide post-exposure prophylaxis (PEP), which is almost 100% effective in preventing the transmission of rabies when administered according to schedule ^8,46^.17.7% more people in urban areas than peri-urban areas visited a health facility after a dog bite.

Distance to the nearest health center is inversely related to receiving medical care for a dog bite. In all of ASA, people who received medical treatment following a dog bite lived significantly closer to a health post or health center than those who did not receive treatment. However, within peri-urban areas, only median shortest distance to a health center (not mean shortest distance to a health center) is associated with receiving medical treatment. These findings suggest that greater distance to nearby health centers or health posts pose a barrier to receiving dog bite treatment. Distance to the nearest health center is a particularly important barrier to medical treatment in the peri-urban areas of ASA, where the average distance to the nearest health center or health post to people bitten by a dog was 2427.2 meters as compared with 448.5 meters in urban areas of the district. Furthermore, newly-developed communities in peri-urban areas are often served by health posts rather than larger health centers. In a previous study of another peri-urban area of Arequipa, focus group participants expressed that disrespect from healthcare professionals at health posts and a shortage of materials like PEP vaccines discourages community members from seeking rabies-related healthcare there ^44^.

These findings align with previous studies, which have reported an association between distance to health facilities and decreased frequency of treatment and prevention for a variety of infectious and non-infectious diseases. ^36,40-42,47-50^. Distance to healthcare facilities is also related to worse health outcomes, especially in rural environments/settings because sick people receive delayed or no medical attention at all ^39,51-58^. Specifically, in the case of rabies, increased distance to a health facility has been associated with a delay in PEP uptake in Dehli and Northwestern Tanzania ^59,60^. In a study of districts throughout Tanzania, there was also a greater number of deaths from rabies the farther away people lived from their closest health facility. Like in other geographic areas, in Arequipa, Peru, people who live farther from the urban center are less likely to receive rabies-related healthcare.

The time required to attend a medical appointment, especially if it is far away, can be a second barrier to receiving PEP and rabies-related medical care. This barrier can be especially salient for people living in peri-urban areas, many of whom work informal jobs ^44,45^, where missing work may mean missing a day’s wages or even putting their employment at risk ^61^. In addition, people who lived in their homes for fewer years are significantly less likely to receive medical follow-up after a dog bite in all of ASA. The same trend was observed in the model stratified by area type, but it was not significant in the peri-urban area most likely due to the reduced sample size. This finding indicates another way in which community members living in peri-urban areas are at a disadvantage in receiving follow-up medical care. People who have lived in their neighborhoods for longer have likely learned the medical and social services available to them and utilize them with more facility. Therefore, a high rate of recent migration in peri-urban neighborhoods could represent another influence on low rates of rabies treatment in peri-urban areas of ASA.

Medical follow-up for dog bites has important public health implications both because PEP during follow-up appointments and because most reports of rabies exposures are made at health centers and health posts ^62^. The difference between the findings from this door-to-door survey and official public health department metrics implies significant underreporting of dog bites, particularly in peri-urban areas of Arequipa. Thus, underreporting is likely due to a number of related barriers, such as distance to the nearest health post, lack of access to well-resourced health posts and disrespect from medical professionals. Despite the fact that no native human rabies cases have been detected in Arequipa to date, around 50 dog rabies cases per year since 2016, low rates of reporting during medical follow-up entails a risk for underdetection of human cases. Missing cases have negative consequences for national programs that control outbreaks and provide preventive measures ^62-66^. We suggest that spatial inequality obscures underreporting, since several factors associated with not receiving medical follow-up or reporting bites are concentrated in peri-urban areas.

Findings should be understood within the context of some limitations. The percentage of people bitten by dogs represents only adults in peri-urban and urban areas of ASA. Children were excluded because adults in the household make decisions about medical treatment. Although we did not analyze children’s data, in the last year, 3.5% (154/4370) of houses interviewed had children bitten by a dog that they did not own. We did not ask about children bitten by dogs that they did own. Since children have made up a disproportionate percentage of dog bite victims ^67-70^ and human rabies cases ^71,72^ in previous studies, future studies of rabies risk in Arequipa should include children to gain a fuller understanding of rabies risk in peri-urban Arequipa. Another limitation is that the data is self-reported in door-to-door surveys. Thus, data may be distorted by participants’ memories or social desirability bias, when participants provide the answer that they think the interviewer or other community members view most favorably.

This study adds to the public health literature by considering rabies specifically in peri-urban communities, areas that have historically been overlooked in academic studies because they do not fit neatly into an urban-rural dichotomy^73,74^. Peri-urban communities face systemic factors that make distance to the nearest health facility a particularly important barrier. The construction of health facilities in newly urbanizing areas often lags behind the population of these areas and community members attend the nearest health facility. Facilities in newly developing areas generally provide basic facilities that offer treatment for frequent health problems ^24^. Initiatives that focus on expanding and modernizing peri-urban health facilities could decrease spatial inequality in rabies risk.

Our study shows that there is substantial underreporting of dog bites throughout Arequipa and particularly in peri-urban areas. We also find that in areas that are just blocks apart, many risk factors for human rabies, such as presence of free-roaming dogs, incidence of dog rabies, dog bite rates, and lack of access to health services, vary greatly. In peri-urban areas of the district we examine, residents have a higher rate of dog bites, likely because of the high density of free-roaming dogs. Then, they are less likely to receive medical follow-up, where wound care and PEP are provided and reporting often occurs. We show that people are more likely to receive follow-up care the closer they are to a medical center or post and discuss how medical infrastructure in peri-urban areas often lag behind urban areas of the city. Therefore, future interventions to address rabies risk in Arequipa should focus on addressing the specific needs of peri-urban areas of the city.

## Data Availability

We have not made available all data referred to in the manuscript. Public availability of geographic coordinates would compromise participants confidentiality and privacy.

## REFERENCES

1. Bayer, A. M. et al. Chagas Disease, Migration and Community Settlement Patterns in Arequipa, Peru. PLoS Negl Trop Dis 3, (2009).

2. Pedersen, D., Tremblay, J., Errázuriz, C. & Gamarra, J. The sequelae of political violence: assessing trauma, suffering and dislocation in the Peruvian highlands. Social science & medicine (1982) 67, 205–217 (2008).

3. Las Migraciones Internas en el Peru: 1981-1993. https://www.inei.gob.pemediaMenuRecursivopublicacionesdigitalesEstLibcapi.htm at <https://www.inei.gob.pe/media/MenuRecursivo/publicaciones_digitales/Est/Lib0018/capi3001.htm >

4. Duranton, G. & Puga, D. in Handbook of Economic Growth Vol II 781–853 (Elsevier, 2014).

5. Levy, M. Z. et al. Urbanization, land tenure security and vector-borne Chagas disease. Proceedings. Biological sciences / The Royal Society 281, 20141003 (2014).

6. Castillo-Neyra, R. et al. Spatial Association of Canine Rabies Outbreak and Ecological Urban Corridors, Arequipa, Peru. Tropical Medicine and Infectious Disease 2, 38 (2017).

7. DGE. Boletín epidemiológico del Perú SE 15. 19 (Centro Nacional de Epidemiología, Prevención y Control de Enfermedades EDITORIAL, 2015).

8. World Health Organisation. WHO Expert Consultation on Rabies. Second report. World Health Organization technical report series 1–139– back cover (2013).

9. DGE. Boletín epidemiológico del Perú SE 52. 40 (Centro Nacional de Epidemiología, Prevención y Control de Enfermedades, 2016).

10. GERSA Arequipa. Boletín epidemiológico semanal Nro 50. 10 (GERENCIA REGIONAL DE SALUD AREQUIPA, 2015).

11. GERSA Arequipa. Boletín epidemiológico semanal Nro 50. 683–695 (GERENCIA REGIONAL DE SALUD AREQUIPA, 2016).

12. DGE. Boletín epidemiológico del Perú SE 1. 24, 22–24 (Centro Nacional de Epidemiología, Prevención y Control de Enfermedades EDITORIAL, 2016).

13. DGE. Boletíin epidemiológico del Perú SE 52. 40 (Centro Nacional de Epidemiología, Prevención y Control de Enfermedades EDITORIAL, 2017).

14. DGE. Boletín epidemiológico del Perú SE 52. 85 (Centro Nacional de Epidemiología, Prevención y Control de Enfermedades, 2018).

15. DGE. Boletín epidemiológico del Perú SE 26. 636–640 (Centro Nacional de Epidemiología, Prevención y Control de Enfermedades EDITORIAL, 2019).

16. Fishbein, D. B. in The Natural History of Rabies (Ed, G. M. B.) 519–549 (CRC Press, 1991).

17. Hemachudha, T., Laothamatas, J. & Rupprecht, C. E. Human rabies: a disease of complex neuropathogenetic mechanisms and diagnostic challenges. The Lancet Neurology 1, 101–109 (2002).

18. Cleaveland, S. & Hampson, K. Rabies elimination research: juxtaposing optimism, pragmatism and realism. Proc. R. Soc. B 284, 20171880 (2017).

19. Hampson, K. et al. Estimating the global burden of endemic canine rabies. PLoS neglected tropical diseases 9, e0003709 (2015).

20. Hampson, K. et al. Transmission dynamics and prospects for the elimination of canine rabies. PLoS biology 7, e53 (2009).

21. Mindekem, R., Lechenne, M. S., Naissengar, K. S. & Lechenne, M. S. Cost Description and Comparative Cost Efficiency of Post-Exposure Prophylaxis and Canine Mass Vaccination against Rabies in N’Djamena, Chad. Front. Vet. Sci. 4, 38 (2017).

22. Rupprecht, C. E., Hanlon, C. A. & Slate, D. Control and prevention of rabies in animals: paradigm shifts. Developments in Biologicals 125, 103–111 (2006).

23. Fooks, A. R. et al. Current status of rabies and prospects for elimination. The Lancet 6736, 1–11 (2014).

24. Ministerio de Salud. Norma Tecnica de Salud para la Vigilancia, Prevención y Control de la Rabia Humana en el Perú. (2017). at <http://bvs.minsa.gob.pe/local/MINSA/4193.pdf>

25. Schneider, M. C. et al. Current status of human rabies transmitted by dogs in Latin America. Cadernos de saúde pública 23, 2049–2063 (2007).

26. Franka, R. et al. Current and future tools for global canine rabies elimination. Antiviral research 100, 220–225 (2013).

27. Tschopp, R., Bekele, S. & Aseffa, A. Dog Demography, Animal Bite Management and Rabies Knowledge-Attitude and Practices in the Awash Basin, Eastern Ethiopia. PLoS Negl Trop Dis 10, 1–14 (2016).

28. Yan, S., Chen, Y., Ye, W., Chen, F. & Li, L. Characteristics and factors associated with post-exposure prophylaxis (PEP) treatment of dog and cat bites among left-behind children : a cross-sectional study in two cities of China. BMJ Open 1–7 (2019). doi:10.1136/bmjopen-2018-024764

29. Ichhpujani, R. L. et al. Knowledge, attitude and practices about animal bites and rabies in general community--a multi-centric study. J Commun Dis 38, 355–361 (2006).

30. Memorial Anual 2017. httpmunialtoselvaalegre.gob.petransparenciainforpresupuestalmemoriaanual.pdf at <http://munialtoselvaalegre.gob.pe/transparencia/infor_presupuestal/memoria_anual_2017.pdf >

31. Manning, S. et al. Human Rabies Prevention --- United States, 2008 cdc.gov (2008). at <https://www.cdc.gov/mmwr/preview/mmwrhtml/rr5703a1.htm>

32. Crickard, P. Leaflet, js essentials. (2014).

33. Liedman, P. Leaflet Routing Machine.

34. Directions API. (2017). at <https://docs.mapbox.com/help/glossary/directions-api/>

35. Bourne, P. A., Eldemire-shearer, D., Mcgrowder, D. & Crawford, T. Examining health status of women in rural, peri-urban and urban areas in Jamaica. N Am J Med Sci. 1, 256–271 (2009).

36. Guagliardo, M. F. Spatial accessibility of primary care : concepts, methods and challenges. International journal of health geographics 3, 1–13 (2004).

37. Kelly, C., Hulme, C., Farragher, T. & Clarke, G. Are differences in travel time or distance to healthcare for adults in global north countries associated with an impact on health outcomes? A systematic review. BMJ Open 6, 1–9 (2016).

38. Kyriopoulos, I. et al. Barriers in access to healthcare services for chronic patients in times of austerity : an empirical approach in Greece. International Journal for Equity in Health 13, 1–7 (2014).

39. Starfield, B., Shi, L. & Macinko, J. Contribution of Primary Care to Health Systems and Health. Milbank Q 83, 457–502 (2005).

40. Castillo-Neyra, R. et al. Barriers to dog rabies vaccination during an urban rabies outbreak: Qualitative findings from Arequipa, Peru. PLoS neglected tropical diseases 11, e0005460 (2017).

41. Fone, D. L., Christie, S. & Lester, N. Comparison of perceived and modeled geographical access to accident and emergency departments : a cross-sectional analysis from the Caerphilly Health and Social Needs Study. International journal of health geographics 5, 1–10 (2006).

42. Jacobs, B., Ir, P., Bigdeli, M., Annear, P. L. & Van Damme, W. Addressing access barriers to health services: an analytical framework for selecting appropriate interventions in low-income Asian countries. Health Policy and Planning 27, 288–300 (2012).

43. Sanchez, R. S. et al. Efecto de la saliva de Triatoma infestans en la duración de la parasitemia de Trypanosoma cruzi en cuyes.

44. Castillo-Neyra, R. et al. Behavioral and structural barriers to human post-exposure prophylaxis and other preventive practices during a canine rabies epidemic. medRxiv 2020.01.16.20016394 (2020). doi:10.1101/2020.01.16.20016394

45. Castillo-Neyra, R. et al. Barriers to dog rabies vaccination during an urban rabies outbreak: Qualitative findings from Arequipa, Peru. PLoS neglected tropical diseases 11, 1–21 (2017).

46. Tarantola, A., Tejiokem, M. C. & Briggs, D. J. Evaluating new rabies post-exposure prophylaxis (PEP) regimens or vaccines. Vaccine 37 Suppl 1, A88–A93 (2019).

47. Jordan, H., Roderick, P., Martin, D. & Barnett, S. Distance, rurality and the need for care : access to health services in South West England. International journal of health geographics 3, 1–9 (2004).

48. O’Donnell, O. Access to health care in developing countries : breaking down demand side barriers. Cad. Saúde Pública 23, 2820–2834 (2007).

49. Turnbull, J., Martin, D., Lattimer, V., Pope, C. & Culliford, D. Does distance matter? Geographical variation in GP out-of-hours service use: an observational study. British Journal of General Practice 471–477 (2008). doi:10.3399/bjgp08X319431

50. Kiwanuka, S. N. et al. Access to and utilisation of health services for the poor in Uganda: a systematic review of available evidence. Transactions of the Royal Society of Tropical Medicine and Hygiene 102, 1067–1074 (2008).

51. Becher, H. et al. Risk factors of infant and child mortality in rural Burkina Faso. Bulletin of the World Health Organization 82, 265–273 (2004).

52. Den Broeck, J. V., Eeckels, R. & Massa, G. Maternal Determinants of Child Survival in a Rural African Community. International journal of epidemiology 25, 998–1004 (1996).

53. Ensor, T. & Cooper, S. in Health, Nutrition and Population (HNP) Discussion Paper (The International Bank for Reconstruction, Development)78 (THE WORLD BANK, 2004).

54. Joshi, R., Jan, S., Wu, Y. & Macmahon, S. Global Inequalities in Access to Cardiovascular Health Care. Our Greatest Challenge. JAAC 52, 1817–1825 (2008).

55. Magnani, R. J. et al. The Impact of Primary Health Care Services on Under-Five Mortality in Rural Niger. International journal of epidemiology 25, 568–577 (1996).

56. Mayxay, M. et al. Respiratory illness healthcare-seeking behavior assessment in the Lao People’s Democratic Republic (Laos). BMC Public Health 13, 12 (2013).

57. Mwaura, L. W., Wandibba, S. & Olungah, C. O. Effect of distance on access to health services among women with type 2 diabetes in a rural community in Kenya. African Journal of Diabetes Medicine 25, 2–4 (2017).

58. Pierce, C. Distance and access to health care for rural women with hearth failure. Online Journal of Rural Nursing and Health Care 7, 27–34 (2007).

59. Joseph, J., n, S., Khan, A. M. & Rajoura, O. P. Determinants of delay in initiating post-exposure prophylaxis for rabies prevention among animal bite cases: hospital based study. Vaccine 32, 74–77 (2013).

60. Hampson, K. et al. Rabies exposures, post-exposure prophylaxis and deaths in a region of endemic canine rabies. PLoS neglected tropical diseases 2, e339 (2008).

61. Machado, R. The informal economy in peru: Magnitude and determinants. Apuntes 41, 191 (2014).

62. Gibbons, C. L. et al. Measuring underreporting and under-ascertainment in infectious disease datasets : a comparison of methods. BMC Public Health 14, 1–17 (2014).

63. Burton, D. C. et al. Healthcare-seeking Behaviour for Common Infectious Disease-related Healthcare-seeking Behaviour for Common Infectious Disease-related Illnesses in Rural Kenya : A Community-based House-to-house Survey. J Health. Popul Nutr 29, 61–70 (2011).

64. Hitchcock, P., Chamberlain, A., Van Wagoner, M., Inglesby, T. V. & O’Toole, T. Challenges to global surveillance and response to infectious disease outbreaks of international importance. Biosecur Bioterror 5, 206–227 (2007).

65. Moss, R., Zarebski, A. E., Carlson, S. J. & McCaw, J. M. Accounting for Healthcare-Seeking Behaviours and Testing Practices in Real-Time Influenza Forecasts. Trop. Med. Infect. Dis. 4, 1–19 (2019).

66. Wójcik, O. P., Brownstein, J. S., Chunara, R. & Johansson, M. A. Public health for the people: participatory infectious disease surveillance in the digital age. Emerging themes in epidemiology 11, 7

67. Dhand, N. K. & Ward, M. P. Human rabies post exposure prophylaxis in Bhutan, 2005-2008: trends and risk factors. Vaccine 29, 4094–4101 (2011).

68. Sudarshan, M. K. et al. An epidemiological study of animal bites in India: results of a WHO sponsored national multi-centric rabies survey. J Commun Dis 38, 32–39 (2006).

69. Kent, S. J. W., Naicker, B. & Wood, D. R. Demographics and management of dog bite victims at a level two hospital in KwaZulu-Natal. S. Afr. Med. J. 102, 845–847 (2012).

70. Kabeta, T., Deresa, B., Tigre, W., Ward, M. P. & Mor, S. M. Knowledge, Attitudes and Practices of Animal Bite Victims Attending an Anti-rabies Health Center in Jimma Town, Ethiopia. PLoS neglected tropical diseases 9, e0003867 (2015).

71. Dodet, B. et al. Rabies awareness in eight Asian countries. Vaccine 26, 6344–6348 (2008).

72. Sriaroon, C., Sriaroon, P., Daviratanasilpa, S., Khawplod, P. & Wilde, H. Retrospective: animal attacks and rabies exposures in Thai children. Travel Med Infect Dis 4, 270–274 (2006).

73. Simon, D. Simon, D. (2008). Urban environments: issues on the peri-urban fringe. Annual review of environment and resources, 33. Annual review of environment and resources 33,

74. Marshall, F., Waldman, L., MacGergor, H., Mehta, L. & Randhawa, P. On the edge of sustainability: perspectives on peri-urban dynamics. (2009).

